# Men’s Internet Sex Addiction Predicts Sexual Objectification of Women Even After Taking Pornography Consumption Frequency into Account

**DOI:** 10.1101/2024.10.12.24315374

**Authors:** Pavla Nováková, Edita Chvojka, Anna Ševčíková, Lukas Blinka, Paul Wright, Steven Kane

## Abstract

Excessive online video pornography consumption is associated with sexual objectification, particularly in male consumers. However, previous studies have not considered that there is a subgroup of internet users whose consumption may become excessive due to their internet sex addiction. Such users may, in response to internet sex addiction symptoms such as craving, have increased levels of sexual objectification. In a sample of 1,272 male consumers of online video pornography (Mage = 32.93, SDage = 9.44), we examined whether internet sex addiction is linked to sexual objectification. We fitted a series of structural equation models and found that men who scored higher on internet sex addiction were more likely to objectify women. More importantly, this link did not cease when controlling for the frequency of online video pornography consumption. Our findings suggest that there are other mechanisms related to addictive symptomatology, than just the link through online video pornography consumption, that may contribute to sexual objectification. Addiction-related factors may have a unique role in fostering sexual objectification. Isolating internet sex addiction as a potential driver highlights the need to address objectifying behaviors in individuals struggling with this addiction.

---

The sexually motivated use of the internet serves nowadays not only to seek sexual pleasure but also to avoid boredom or reduce stress (Böthe et al., 2021). Although people may search for information about sex, read erotic stories, have sex over a webcam or chat, search for sexual partners, and browse online sex shops (Ballester-Arnal et al., 2021), online video pornography (further just “pornography”) consumption remains currently the most prevalent activity (Wright et al., 2023; Hoagland & Grubbs, 2021), especially among male internet users.

Outcomes of frequent exposure to nudity and sexual activities (Campbell & Kohut, 2017) have been subject to a significant body of research (see Peter & Valkenburg, 2016; Harkness et al., 2015). Studies on the perceived positive effects of pornography consumption involve positive links with greater openness towards sexual experimentation, improved communication about sex, heightened sexual comfort, the exploration of sexual interests, understanding one’s sexuality, and better knowledge of anatomy, physiology, and sexual behaviour (Kohut et al., 2017; Hesse & Pedersen, 2017; Castro-Calvo et al., 2018). However, pornography consumption has also been researched in connection to adverse consequences, ranging from developing unhealthy sexual behaviour, such as internet sex addiction, to acquiring problematic sexual beliefs and attitudes toward the opposite gender that pornography content may mediate and communicate (Baltazar et al., 2010; Harper & Hodgins, 2016; Herbenick et al., 2020; Lim et al., 2016; Paslakis et al., 2022; Wright et al., 2022).

Substantial research work has shown the linkage between frequent pornography exposure and sexual objectification, particularly the treatment of women as sex objects (Willis et al., 2022; Wright & Tokunaga, 2016). While objectified individuals in pornography are not limited to women but also include men (Bridges et al., 2024) and people beyond the gender binary (Pavanello, 2023), we expand the research on objectification of women for several reasons grounded in the current literature. Research has consistently shown that pornography typically emphasises male pleasure, with a typical script focusing on male orgasms and close-ups of female bodies (Klaassen & Peter, 2015), which further reinforces the objectification of women. Furthermore, men who consume online pornography more frequently are more likely to perceive women as sex objects (Bridges et al., 2024; Wright & Tokunaga, 2016; Peter & Valkenburg, 2009; Vandenbosch & Oosten, 2017). This relationship is especially pronounced for male consumers, likely because pornographic content is often tailored toward male preferences (Miller & McBain, 2022) and because men tend to watch these videos more often and more intensively than women (Solano et al., 2018; Campbell & Kohut, 2017).

Frequent pornography use is not only typical behaviour for non-problematic pornography users but also for individuals at risk of internet sex addiction (Böthe et al., 2020; Grubbs et al., 2018). Despite the growing body of research on sexual objectification, no studies to date have explored whether the sexual objectification exhibited by sex-addicted internet users exclusively relates to frequent pornography use or if it broadly connects to the symptoms of internet sex addiction. This pioneering research aims to examine whether internet sex addiction uniquely contributes to sexual objectification, even when accounting for the frequency of pornography use. By exploring the topic of sexual objectification, we hope to contribute towards a more nuanced understanding of individuals experiencing symptoms of internet sexual addiction.

## Sexual Objectification

Sexual objectification puts the physical appearance or sexual functionality of the body above the overall value of the person (e.g., Fredrickson & Roberts, 1997; Szymanski et al., 2011). An essential dimension of sexual objectification is the treatment of a person as a body (or its parts) whose only purpose is to provide sexual pleasure (Bartky, 2015). Some feminists and psychologists (Daniels et al., 2020) believe that sexual objectification negatively shapes women’s self-image since it makes society neglect their competence and intelligence. Sexual objectification can also lead to adverse psychological effects, including depression, sexual dysfunction, or eating disorders (Ward et al., 2023). Although both men, women, and beyond binary-gendered people can be targets of sexual objectification, the concept is strongly associated with women (Rodríguez-Castro et al., 2018). Compared to men, images of women are more likely to display heightened attention to the body parts than the whole body (Gervais et al. (2012).

Many channels objectify women (e.g., TV programs, video games, and online media). However, one with the most potent effect is pornography (Willis et al., 2022). Klassen and Peter (2015) concluded that women in pornographic material are more likely to be depicted in a reductive sense. A higher frequency of pornography consumption relates to a higher level of sexual objectification. Wright and Tokunaga (2016) found that the frequency of men’s exposure to online video pornography significantly correlated with objectified cognitions about women (which also predicted stronger attitudes supportive of violence against women). Vandenbosch and Eggermont (2019) revealed that adolescents are more likely to believe that women are sex objects if they consume sexually explicit internet materials more often. Pornography may, therefore, convey the acceptability of treating people as sexual objects.

## Excessive Online Sexual Activities

Some authors claim that online addictions have become a new risk for mental health (Venkataramu et al., 2021). According to Yau et al. (2013), at-risk/problematic internet users are more likely to score poorly on measures of self-control, impulsivity, and depression. Consequently, such addictive behaviour strongly negatively correlates with various subjective health outcomes (Böthe et al., 2021; Kraus et al., 2016; Purwaningsih & Nurmala, 2021).

Excessive use of online materials has been conceptualised as a phenomenon that may fall within the hypersexual, compulsive-impulsive, or addictive spectrums of disorders. However, only Compulsive Sexual Behaviour Disorder, which includes problematic pornography use, has been determined to be an official diagnosis (ICD-11, 2019), with internet sex addiction as a subcategory (Fuss et al., 2019; Sassover & Weinstein, 2022). Nevertheless, there is a growing number of studies according to which the excessive use of the internet for sexual purposes may be a sign of behavioural addiction (Gola et al., 2017; Karila et al., 2014; Blinka et al., 2022). Authors supporting this thesis (Cooper et al., 2000; Schwartz & Southern, 2013; Orzack & Ross, 2000; Schneider, 2000) argue that with the accessibility of pornography and other online sexual content, some internet users fail to control such consumption to the extent that they fulfil the behavioural addiction criteria.

These criteria within the addiction model are salience, loss of control, relapse, negative consequences (e.g., conflicts and problems), tolerance, withdrawal symptoms, and craving (Griffiths, 2012; Karila et al., 2014; Wölfling et al., 2012). Until now, the excessive use of the internet for sexual purposes was, according to the Compulsive Model, considered a tool to decrease anxiety, implying that the last criterion, craving, does not fit into this model. However, Blinka et al. (2022) and other researchers (Chen et al., 2018) show that pleasure may be the trigger that drives internet users, including sex-addicted people, to search for sexual stimuli (e.g. intentional exposure to sexual objects).

## The Mechanisms Behind the Connection Between Online Video Pornography Consumption, Internet Sex Addiction and Sexual Objectification

Incorporating a variety of media effects, behavioural theories, and information-processing theories (e.g., Bandura, 2014; Brown, 2002), the sexual script acquisition, activation, application model (3 AM) explains how and under what circumstances consuming pornography affects sexual cognition, including beliefs about sexual objectification (Wright, 2020; Wright & Tokunaga, 2016; Vandenbosch & van Oosten, 2017). Wright’s sexual script acquisition, activation, and application model (3 AM; Wright, 2011) integrates the sexual script theory (Simon & Gagnon, 2003) and Huesmann’s (1986) reasoning about mass media as a cognitive script provider. The 3 AM implies that pornography may modify consumers’ sexual cognition, including beliefs about sexual objectification, through the following mechanisms. Pornography users become exposed to scripts they are not aware of (acquisition) and primed with scripts of which they are already aware (activation). Moreover, pornography encourages the utilisation of such scripts by portraying specific sexual behaviours or patterns of sexual behaviour as rewarding, appropriate, and normative (application). Due to the focus of mainstream pornography on primarily male audiences, especially male pornography consumers may start following salient sexual scripts that may be of seemingly functional value for them, such as the scripts portraying women as tools for male pleasure. The functional values and saliency of online pornographic material may then support and sustain the frequent exposure. Frequent exposure is thus likely to enhance long-term memory activation and make sex-objectifying scripts more accessible in situations when frequent pornography users evaluate their perception of and attitudes toward women (Wright, 2019).

Sexual objectification might not only be a consequence of pornography consumption, as suggested by the 3 AM. In addicted people, objectification can also be maintained and strengthened as a response to cravings and experiences related to withdrawal from internet use and access to sexually explicit stimuli. Recent studies suggest that viewing women as sexual objects helps sexually addicted people cope with their irritation and craving from their lack of access to online pornography (Blinka et al., 2022; Ševčíková et al., 2018; Grubbs et al., 2019). In Blinka et al., participants acknowledged their tendency to *‘exploit sexual objects visually’* (i.e., to create a mental image full of sexual stimuli) when they did not have access to online pornography. The participants confided that, when abstaining, they were irritated by the lack of access to pornographic images to the extent that they tried to draw as much as possible from each potential sexual object (either in their environment or activated in their memory) to nourish their fantasy. Sexually addicted men may adopt specific behavioural actions, such as walking near a nudist beach or leering at women from a balcony, to relieve or avoid withdrawal symptoms. According to Ševčíková et al., this behavioural pattern, which is based on the extensive sexual objectification of women, seems to saturate mental content with sexual stimuli when watching pornography is avoided or not available. Moreover, addicted persons include this behavioural pattern in the list of activities they should avoid.

In this respect, we propose to test the following hypotheses:

**H1:** Internet sex addiction significantly predicts sexual objectification with a small/medium positive effect.
**H2:** The relationship between internet sex addiction and sexual objectification remains significant even after controlling for online video pornography consumption frequency.

## Materials and Methods

### Data Collection and Participants

The data come from a convenience sample online survey about internet sex conducted in the Czech Republic in 2017. The study was primarily advertised on the biggest Czech online erotic platform, Amateri.com (2,215 participants, which is 88% of the whole sample, came from this website) and via social networking sites that targeted respondents who engaged in online sexual activities. A total of 2,518 participants aged 18 to 77 (M_age_ = 32.73, SD_age_ = 9.62; 73.2% men) filled in the questionnaire in the Czech language. For this study, we worked with a subset of 1,272 men (M_age_ = 32.93, SD_age_ = 9.44) who self-identified as heterosexual, who filled in more than 75% of the questionnaire, and whose age distribution aligned with the whole sample. The choice of 75% was arbitrary, but it reflected our motivation to avoid the inclusion of careless, bored, or otherwise demotivated participants and related response biases. There were 1,312 non-homosexual men in total; no man indicated bisexuality.

The data collection occurred from April to November 2017 on the Lime Survey platform. The participants were informed about the purpose of the study, data management and analytical procedures, and their rights. This study was conducted in line with the university’s ethical guidelines. Details about the study aim, procedures, data collected, and the minimum age (18 years old) were provided on the first page of the questionnaire. Participation was voluntary and all information provided was confidential. We did not collect any personal data, and no incentives were offered to participants. A written informed consent was also obtained.

### Measures

#### Internet Sex Addiction

Internet Sex addiction in the preceding 12 months was measured with the Short French Internet Addiction Test Adapted to Online Sexual Activities (s-IAT-sex; Wéry et al., 2016). The scale assesses the potential symptoms of addiction to sexual websites. Wéry et al. developed the instrument to adapt the general IAT (Pawlikowski et al., 2013). The Czech version used in this survey underwent a 3-way back-translation process. The tool consists of 12 items with a 5-point Likert scale that ranges from “never” (1) through “rarely”, “sometimes”, “often” to “very often” (5). The items tap into the core criteria of addiction. The scale has two dimensions: (1) the loss of control and time management: e.g., “How often do you find that you stay on internet sex sites longer than you intended?” and (2) craving and social problems: e.g., “How often do you choose to spend more time on internet sex sites than going out with others?”)) measured by six items each. According to Wéry et al., the tool shows good psychometric properties. However, the correlation between the two latent factors in the original study was extremely high (0.89), and recent literature, including the authors themselves (e.g., Chen & Jiang, 2020; Levi et al., 2020; Wéry et al., 2018, 2020) has exclusively treated s-IAT-sex as unidimensional. This opens the question of whether the two factors represent distinct dimensions. The distributions of most s-IAT-sex items were notably skewed (skewness: mean = 1.33, median = 1.37, min = -0.04, max = 2,52; kurtosis: mean = 1.61, median = 1.29, min = -0.77, max = 6,39). This indicates that s-IAT-sex would prevent the pathologising of internet use for sexual purposes but, consequently, was not able to discriminate between the different levels of addiction severity. In alignment with the literature, we also treated the scale as unidimensional (and elaborated on our motivation for this decision in the Results section). The reliability of the general factor was high (McDonald’s ω_total_ of .89).

#### Sexual objectification

Sexual objectification was measured by five items (full version in Appendix) that addressed the frequency of typical objectifying behaviour (e.g., undressing passerbyes with eyes). The respondents indicated the frequency of such behaviour with a 5-point Likert scale, going from “never” through “rarely”, “sometimes”, “often” to “very often”. The item wording was based on semi-structured interviews with sexually addicted men who searched for treatment (Anonymised source, XXXX). The responses were fairly normally distributed (skewness: mean = -0.09, median = 0.09, min = -0.68, max = 0.23; kurtosis: mean = -0.49, median = -0.52, min = -0.83, max = 0,12). We treated sexual objectification as a latent trait for our analysis and estimated a one-factor model over the five items. The measure’s reliability was high (McDonald’s ω_total_ of .86).

#### The Frequency of Online Video Pornography Consumption

Consumption frequency was captured by one item: “How often have you used the internet to watch online video pornography in the last 12 months?” The participants answered on a 7-point Likert scale going from “daily” (=1) through “two times to five times a week”, “once a week”, “two times or three times a month”, “once a month”, “less than once a month” to “not once” (=7), and the item was re-coded so that (1) would indicate “not once” and (7) would indicate “daily”. The responses were fairly skewed (skewness = -1.52, kurtosis = 2.52).

### Data cleaning and preparation

We started with the general sample. We excluded all women and homosexual men. We then excluded people with more than 25% missing responses. Subsequently, we imputed the missing data points with the CART method (van Buuren & Groothuis-Oudshoorn, 2011), which is a procedure with an origin in machine learning that is based on classification trees that perform well with heavily skewed data (Burgette & Reiter, 2010). We then performed a non-paranormal transformation to move the data distribution closer to normal (*gaussianization*, Jiang et al., 2021). We checked for outliers by visually inspecting boxplots and performing Grubb’s tests (Komsta, 2022) and found none. This left 1,272 men. We re-ran the preregistered sensitivity analysis (Moshagen, 2021) because the resulting sample was smaller than expected. The analysis indicated a power of ∼1 to detect a small effect.

### Data Analysis

The analysis was pre-registered via the Open Science Framework ([OSF - unblinded](https://osf.io/8hwsq/?view_only=415ca79c6f674aa3ad11b4c6bba74ed); https://osf.io/n4rvb). The analyst had not seen the data beyond the “variable view” in SPSS before downloading them once the registration was approved. The full dataset is not publicly available per the PI’s decision. However, a subset of the data used for this study is available in the associated OSF repository, with the analytic script and a basic codebook. All analyses were conducted in R version 4.2.1 (R Core Team, 2019) and used the lavaan package (Rosseel, 2012).

We used Structural Equation Modeling to test our hypotheses. Since gaussianization was ineffective in transforming the heavily skewed variables, we used a robust weighted least square mean and variance-adjusted estimation method (WLSMV) and polychoric correlations as input. We identified all models by setting latent variances to one. We first established the measurement models by conducting CFAs. Then, we fitted Model 1 with only one direct regression path from Internet sexual addiction to Sexual Objectification. Then, we fitted Model 2 with an additional indirect path via the frequency of online video pornography consumption. Model 1 was nested in Model 2 as it included a subset of the parameters of Model 2. Thus, we conducted a χ^2^ difference test to assess whether adding the indirect path significantly improved fit.

### Outcome Measures

#### χ2 difference test

Since Model 1 is nested in Model 2, we subtracted the two χ2 statistics and the models’ degrees of freedom from each other and reported the significance of this difference. We flagged the model with the lower χ^2^ as better fitting in case of a significant difference.

#### Goodness-of-Fit Indices

Next to the χ^2^ statistic, we report RMSEA and its 95% CI, CFI, TLI, and SRMR. We pre-registered the following criteria to flag a good-fitting model: RMSEA < 0.08, CFI and TLI > 0.9, and SRMR < 0.1. Nevertheless, it is worth mentioning that the conventional goodness-of-fit guidelines, such as those by Hu and Bentler (1999), have been criticised for their lack of generalizability, as their values are sensitive to other model characteristics than the amount of misspecification (see McNeish, 2023; McNeish & Wolf, 2023; Fan & Sivo, 2007). We, therefore, interpret the fit indices only as a rough indication.

#### Bootstrap Confidence Intervals

We also report the 95% bootstrap confidence around the regression coefficients over 1000 resamples to assess the significance of the direct and indirect effects. Should the CIs contain zero, we deem the regression coefficient non-significant.

## Results

### Descriptive Statistics

Descriptive statistics after multiple imputation and gaussianization can be found in Appendix A.

### Measurement Models

We first estimated a model that treated internet sex addiction as a second-order factor over the two dimensions (*loss of control/time management* and *craving/social problems*), as defined by Wéry et al. (2016). While this model fitted the data well, none of the loadings in the loss of control/time management dimension was significant (see Appendix B). Since discarding all the items with non-significant loadings would lead to omitting the original *loss of control/time management*dimension, we decided to deviate from the pre-registration and estimate an alternative model. We first fixed both paths from the second-order factor to the first-order factors to 1, estimating a model equivalent to a two-factor solution with correlated factors. This remedied the problem with the non-significant loadings. However, the resulting model did not fit the data (see Table 1b and Appendix C). Finally, we discarded the second-order factor and fitted a model with one general factor of internet sex addiction, which loaded on all items of s-IAT-sex. This model fitted the data satisfactorily (see Table 1a) and showed substantial factor loadings (ranging from .57 to .83, mean = median loading = .69). We used the one-factor measurement model for testing our hypotheses.

**Table 1.**

|  | mean | sd | median | min | max | skewness | kurtosis |
| --- | --- | --- | --- | --- | --- | --- | --- |
| age | 32,92 | 9,44 | 31 | 18 | 77 | 0,88 | 0,89 |
| frequency of online pornography consumption | 5,84 | 1,22 | 6 | 1 | 7 | -1,52 | 2,52 |
| Internet sex addiction S-IAT-SEX |  |  |  |  |  |  |  |
| 1C Covers visiting sex pages | 1,81 | 1,11 | 1 | 1 | 5 | 1,28 | 0,74 |
| 2C Angry when disturbed from sex pages | 1,36 | 0,79 | 1 | 1 | 5 | 2,52 | 6,39 |
| 3C Thinks of sex activities on internet | 1,86 | 0,92 | 2 | 1 | 5 | 0,89 | 0,13 |
| 4C Hides the amount of time spent on sex pages | 1,70 | 1,12 | 1 | 1 | 5 | 1,59 | 1,46 |
| 5C Does not go out because of sex pages | 1,66 | 0,94 | 1 | 1 | 5 | 1,41 | 1,45 |
| 6C Does not feel himself without internet | 1,37 | 0,76 | 1 | 1 | 5 | 2,31 | 5,32 |
| 1L Visits sex pages for longer than intended | 2,80 | 1,08 | 3 | 1 | 5 | -0,04 | -0,77 |
| 2L Neglects household | 1,69 | 0,93 | 1 | 1 | 5 | 1,32 | 1,20 |
| 3L Limited work performance | 1,50 | 0,81 | 1 | 1 | 5 | 1,71 | 2,61 |
| 4L Limited sleep because of sex pages | 1,98 | 1,04 | 2 | 1 | 5 | 0,86 | 0,00 |
| 5L Wants to stay 'just a few more minutes' | 2,12 | 1,15 | 2 | 1 | 5 | 0,69 | -0,57 |
| 6L Unsuccessful at limiting sex pages | 1,59 | 0,89 | 1 | 1 | 5 | 1,46 | 1,38 |
| Objectification |  |  |  |  |  |  |  |
| 1 Scans someone's figure with his eyes | 3,93 | 0,91 | 4 | 1 | 5 | -0,68 | 0,12 |
| 2 Creepy gaze | 2,77 | 1,13 | 3 | 1 | 5 | 0,11 | -0,73 |
| 3 Observes select bodyparts | 3,30 | 1,02 | 3 | 1 | 5 | -0,20 | -0,49 |
| 4 Observes certain bodyparts instead of listening | 2,66 | 1,05 | 3 | 1 | 5 | 0,23 | -0,52 |
| 5 Undressess passer-byes with his eyes | 2,86 | 1,18 | 3 | 1 | 5 | 0,09 | -0,83 |

*Descriptive statistics before multiple imputation and transformation.* Note. Items labeled “L” originally fall to the loss of control/time management factor, items labeled “C” originally fall into the craving/social problems factor of S-IAT-SEX (Wéry et al., 2016)

**Table 2.** Model Fit and comparison.

| | $\chi^2$ | $p(\chi^2)$ | df | RMSEA<br>90% CI<br>lower | RMSEA | RMSEA<br>90% CI<br>upper | CFI | TLI | SRMR |
| --- | --- | --- | --- | --- | --- | --- | --- | --- | --- |
| Hypothesis testing |  |  |  |  |  |  |  |  |  |
| Model 1: only direct path | 1159,32 | <0,01 | 135 | 0,07 | 0,08 | 0,08 | 0,98 | 0,97 | 0,08 |
| Model 2: direct + indirect path | 558,09 | <0,01 | 133 | 0,05 | 0,05 | 0,05 | 0,99 | 0,99 | 0,05 |
| difference | 128,73 | 2 | <0,01 |  |  |  |  |  |  |
| Measurement models |  |  |  |  |  |  |  |  |  |
| s-IAT-sex 2<sup>nd</sup> order | 503,01 | <0,01 | 116 | 0,05 | 0,05 | 0,06 | 0,99 | 0,99 | 0,05 |
| s-IAT-sex 2-factor | 2515,88 | <0,01 | 118 | 0,12 | 0,13 | 0,14 | 0,94 | 0,93 | 0,11 |
| s-IAT-sex 1-factor | 538,57 | <0,01 | 118 | 0,05 | 0,05 | 0,06 | 0,99 | 0,99 | 0,05 |
| objectification | 36,82 | <0,01 | 5 | 0,05 | 0,07 | 0,09 | 0,99 | 0,99 | 0,03 |

*Notes.* 2nd order = second-order model with two first-order factors based on Wéry et al. (2016); 2-factor = model equivalent to a two-factor solution with correlated factors based on Wéry et al. (2016).

The one-factor model of Sexual Objectification fitted the data well (see Table 1a) and showed substantial factor loadings (ranging from .61 to .76 mean = median loading = .70).

### Hypothesis Testing

The two models are compared in Table 1b and Figure 1a. Model 2 (with the indirect path *internet sex addiction -> frequency -> sexual objectification* added on top of the direct path *internet sex addiction -> sexual objectification*) fitted the data significantly better than Model 1 (only the direct path *internet sex addiction -> sexual objectification*). Model 2 also fitted the data well in terms of the fit measures. Moreover, opening the indirect path via the frequency of consumption did not substantively weaken the direct (moderately strong) relationship between internet sex addiction and sexual objectification. Internet sex addiction predicted the frequency of consumption with a small effect, and the frequency of consumption predicted sexual objectification with a medium-sized effect. Thus, we supported both our hypotheses. A complete overview of all the estimated parameters is in Supplementary Materials (Appendix D).

**Figure 1.**
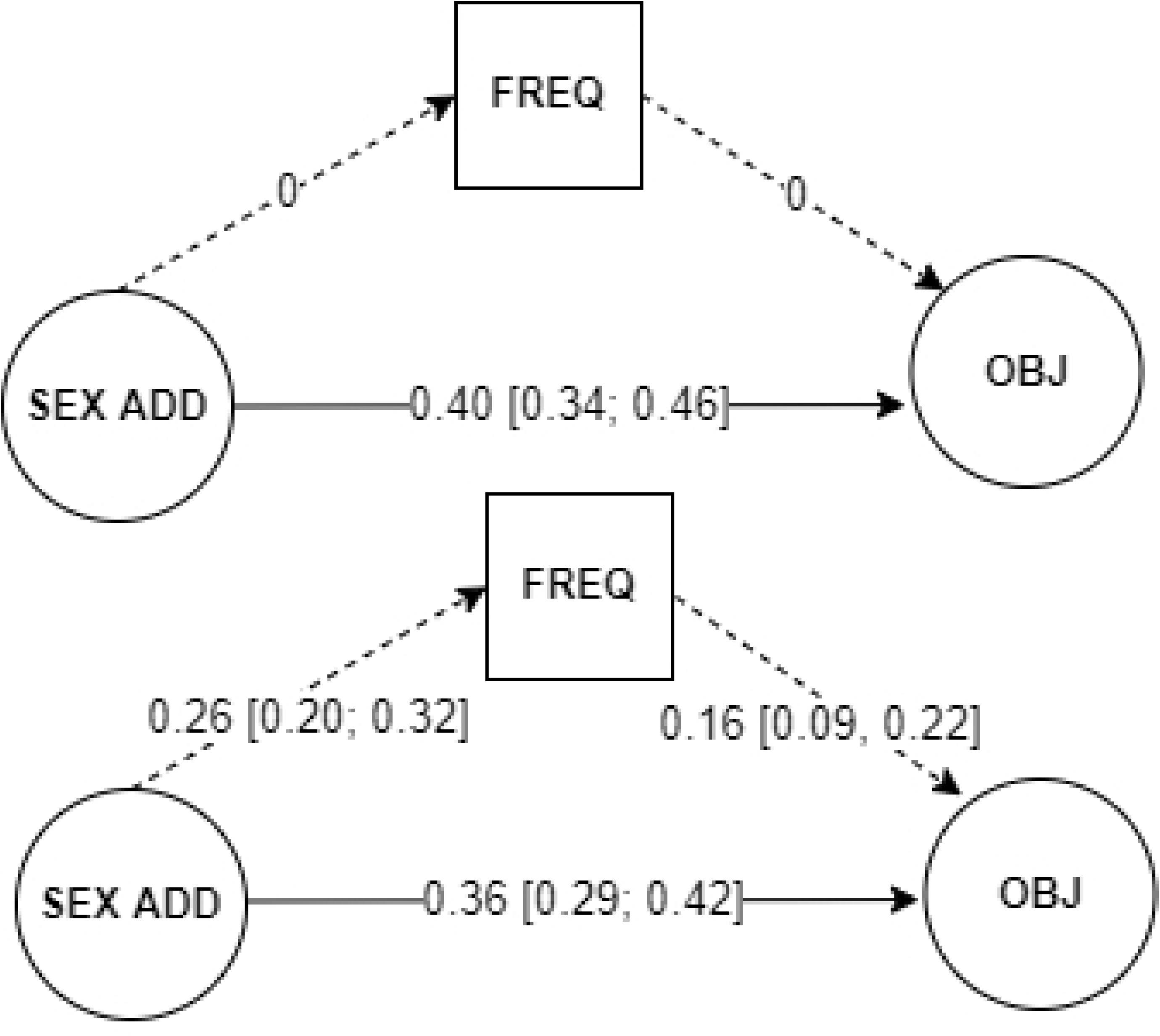

## Discussion

The primary aim of this study was to investigate whether the relationship between internet sex addiction and sexual objectification is more than simply the outcome of frequent exposure to online video pornography. In a sample of heterosexual men who used the internet for sexual purposes, we found a relationship between internet sex addiction and sexual objectification (SO). This direct relationship remained virtually unchanged even after opening the indirect path between online sex addiction and sexual objectification through the frequency of pornography consumption, suggesting that internet sex addiction is directly associated with sexual objectification, regardless of the frequency of consumption.

Our findings contribute to the current understanding by suggesting that sexual objectification is not solely a result of frequent pornography consumption (as proposed by the 3AM, Wright, 2011). Notably, we found a positive relationship between sex addiction and tendencies toward sexual objectification, even after accounting for the frequency of pornography use. In particular, there is a solid direct link between sex addiction and sexual objectification. This finding is in line with prior research according to which sexually addicted men experienced intense urges to exploit sexual objects (i.e., women) visually when they did not either purposefully or unintentionally have access to pornography (Blinka et al., 2022; Ševčíková et al., 2018).

Ševčíková et al. suggest that, in this context, extensive sexual objectification may function as a withdrawal-related behaviour to cope with craving when pornography is avoided or inaccessible.

This extensive sexual objectification allows sexually addicted people to saturate mental content with sexual stimuli to address sexual urges.

The direct link between internet sex addiction and sexual objectification may also point towards other mechanisms that connect internet sex addiction and sexual objectification and, therefore, explain the direct relationship. One such mechanism is the reward-seeking behaviour commonly associated with addiction. In line with the incentive-sensitisation theory (Robinson & Berridge, 2008), individuals with sexual addiction may develop an intensified response to sexual stimuli due to their brain’s heightened sensitivity to these cues. This increased sensitivity makes sexual stimuli more salient, leading to stronger reactions to sexual content (Love et al., 2015). Consequently, sexually addicted men are more likely to engage in sexual objectification, reinforcing this behaviour over time (Chau et al., 2024). Heightened sensitivity to cues could be activated by cravings or withdrawal symptoms (Voon et al., 2014), which may drive objectification even in the absence of active pornography consumption.

Apart from the main findings, this study has two additional methodological outcomes. First, the 5-item scale for sexual objectification developed for the present study showed desirable properties in terms of high and significant factor loadings and high internal consistency. After more thorough construct validation, the short scale can be adopted by other researchers and aid future research on sexual objectification. Second, our analyses did not support the two dimensions of s-IAT-sex as proposed by Wéry et al. (2016). This finding aligns with other studies (e.g., Chen & Jiang, 2020). The original work of Wéry et al. reported a robust correlation between the two dimensions, leading to whether it is plausible to expect the s-IAT-sex instrument to fit a two-dimensional model. In this respect, we do not attribute the unsatisfactory behaviour of two-dimensional models to possible cultural differences but rather to the model itself. Treating the measure (and the construct) as two-dimensional could have severe ramifications if it repeatedly proves problematic across multiple studies. Therefore, we encourage other researchers to revisit the theory behind s-IAT-sex and advocate for treating s-IAT-sex as unidimensional until an alternative dimensional structure is proposed. The difficulties with this two-factor model may suggest that further refinement is needed, potentially integrating the cognitive-behavioural model of pathological use (Davis, 2001), which separates cognitive preoccupation from behavioural consequences. This model may offer additional insights into how sex addiction manifests concerning objectification tendencies.

Our study is not without limitations. Since we only collected cross-sectional data, the causal pattern behind the observed link must be investigated in future studies. Sexual objectification in the form of internalised gendered norms may be a precursor for internet sex addiction. The two constructs can also reciprocally reinforce each other, creating a feedback loop. The sexual objectification measure constructed for this study was undirected concerning different forms of sexual objectification, as well as the gender identity of the objectified. Different forms of sexual objectification, ranging from visual to verbal to behavioural objectification, may operate in various ways depending on the level of addiction severity. Future studies could examine whether specific types of objectification are more strongly linked to addiction symptomatology, adding granularity to the understanding of how addiction and objectification interact.

Moreover, men who self-identify as heterosexual do not exclusively engage with heterosexual pornography (Sharkey et al., 2022) and may also objectify people of other genders (Engeln-Maddox et al., 2011). Since our measure did not capture these nuances, the relationships may be biased when only women are the target of sexual objectification. We worked with a robust, convenient sample of internet users who visited sexually oriented websites but experienced little to no symptoms of internet sex addiction. Those with intense addiction symptomatology may refrain from visiting pornographic sites to self-cure or may be less open about their problematic sexuality. Thus, future studies may find differences in clinical samples. Lastly, although the data were collected in 2017, the study addresses phenomena such as internet sex addiction, pornography use, and sexual objectification, which remain pressing issues even today.

In conclusion, this research highlights that addiction-related symptomatology may have a unique role in fostering sexual objectification. Isolating internet sex addiction as a potential driver may expand the scope of inquiry in the field of sexual behaviour and objectification and highlight the need to address objectifying behaviours in individuals struggling with internet sex addiction. Our findings indicate a need for future studies to explore how addiction-related mechanisms, such as cognitive distortions and cue sensitivity, may contribute to sexual objectification tendencies directed at women and also men and individuals beyond the gender binary. Ultimately, such findings could have vast implications for clinical interventions and broader societal discussions around media consumption and gender relations among diverse internet users.

## Work contribution

Pavla Nováková, Edita Chvojka and Anna Ševčíková contributed to this work by writing the initial draft, reviewing and working on the methodology and analysis. Lukas Blinka participated by collecting the data as well as reviewing. Paul Wright and Steven Kane commented the paper and reviewed the language and grammar.

## Ethics and Publication Malpractice

The authors conform to the Ethics and Publication Malpractice standards set by the Committee on Publication Ethics (COPE). Ethical Committee statement is included in the Attachments.

## Data availability

https://osf.io/n4rvb

## Online supplementary material

https://osf.io/8hwsq

## Blinded supplement

https://osf.io/8hwsq/?view_only=415ca79c6f674aa3ad11b4c6bba74ede

